# Assessment of long-term damage and cardiovascular risk in juvenile systemic lupus erythematosus compared to juvenile dermatomyositis in adulthood

**DOI:** 10.64898/2026.03.30.26349504

**Authors:** Jiagning Li, Imaan Ali, Tharuni Mailoo, Shreya Doddi, Niveda Raj, Edward Palmer, Coziana Ciurtin

## Abstract

**Objectives:** Juvenile systemic lupus erythematosus (JSLE) and juvenile dermatomyositis (JDM) are systemic autoimmune rheumatic diseases (RMDs) with childhood-onset associated with increased risk of damage accumulation and cardiovascular disease (CVD) over the life course.

**Methods:** Damage associated with JSLE and JDM has been assessed using validated outcome measures in a longitudinal single-centre cohort study with long-term follow-up, involving data collected both retrospectively and prospectively. Descriptive statistics, sensitivity and regression analyses have been used to evaluate predictors of damage and CVD-risk.

**Results:** We assessed comparatively a JSLE cohort (n=76), with a mean age of 24.3 ±4.2 years and a JDM cohort (n=79) with a mean 20.1 ± 5.0 years (p<0.001), with matched duration of follow-up (10.0 ± 4.2 vs. 11.0 ± 5.1, respectively, p=0.68). Traditional CVD-risk factors, including hypertension (p=0.02), dyslipidaemia (p=0.0005), and higher total cholesterol (p=0.01) and LDL-cholesterol (p=0.02) levels at the last assessment were higher in JSLE vs. JDM. Over the disease course, 39 (51.3%) AYA with JSLE vs. 47 (59.4%) AYA with JDM accumulated damage (p=0.307), which was independently predicted by the body mass index in both cohorts (p=0.038 and p=0.026, respectively).

The PDAY score was the only tool able to stratify AYA based on CVD-risk (median = 5 (4-13) points in JSLE vs. 0 (0-3) points in JDM, p=0.0001), as all the adult CVD-risk scores were very low in both cohorts.

**Conclusions:** This is the first comparative evaluation of JSLE vs. JDM in adulthood, which highlighted increased damage burden and CVD-risk in JSLE that warrants further investigation.

**Statement of clinical significance:** No previous studies explored log-term outcomes and cardiovascular risk assessment in juvenile systemic lupus erythematosus (JSLE) compared to juvenile dermatomyositis (JDM), despite both conditions sharing pathogenic mechanisms, a significant risk of organ damage, and overlapping therapeutic approaches. Additionally, there is a scarcity of studies following children and young people into adulthood and none evaluating the performance of validated cardiovascular disease (CVD)-risk scores in these two conditions. This study provides the first comparative evaluation of organ specific damage accrual and their predictors, and CVD-risk drivers in JSLE vs. JDM in two cohorts with similar mean disease duration of over 10 years. Our results highlight similar prevalence of overall damage in both conditions, but with distinct organ and systems involvement, and significantly increased CVD-risk in JSLE. Interestingly, in addition to recognised predictors, the body mass index (BMI) was an independent predictor of damage in both cohorts, which emphasizes the need for better strategies to address modifiable contributors to damage accrual. This study raises awareness that CVD-risk scores validated for use in adult populations underperform in both JSLE and JDM in young adulthood, highlighting in parallel the potential clinical utility of a paediatric CVD-risk score (the PDAY score) for risk stratification in JSLE, as well as the need for more robust CVD-risk tools for adequate risk identification and tailored management strategies in JSLE and JDM later in life.

## Introduction

Childhood onset systemic rheumatic diseases (RMDs) are chronic inflammatory conditions associated with dysregulation of the immune system, which start before the age of 18 years. Many RMDs affect multiple organs and systems and are potentially associated with long-term damage, which is particularly relevant for children and young people (CYP) who live with these conditions for longer. RMDs are usually managed with immunosuppression medication, including glucocorticoids (GC), which can increase the risk of infections or cause various side-effects and contribute to damage accrual (1). Additionally, RMD-related chronic inflammation and some of the therapies used in RMDs can increase the cardiovascular disease (CVD)-risk of an individual (2).

Systemic RMDs could have multidimensional impact on general health, functional level, as well as education, social participation and mental health of a young person, with many RMDs having a variable and unpredictable course. Juvenile systemic lupus erythematosus (JSLE) and juvenile dermatomyositis (JDM) are two of the RMDs associated with the most severe and life-threatening presentations in paediatric rheumatology (3). JSLE and JDM share molecular pathogenesis features, especially related to abnormal activation of the interferon (IFN) pathway and mitochondrial dysfunction (4-6), lipid profile dysregulation leading to a pro-atherogenic profile (7, 8), and subsequently, to increased CVD-risk (9-11). Living with a chronic condition with unpredictable course during adolescence poses additional challenges in terms of ensuring optimal disease control and adherence to treatment recommendations (12), on the background of physiological and neurocognitive changes that occur around puberty and adolescence with impact on risk-taking behaviours (13), physical activity (14), diet (15) and CVD-risk profile (2).

In the large UK JSLE cohort (N=430) followed-up for a median duration of only 2 years, 29% CYP accrued damage, which was associated with the disease activity trajectory, with low disease activity states protecting against damage accumulation (16). In a single-centre JDM cohort with 16.8 year follow-up, 90% of young people accumulated damage in at least one domain after a mean of 4.2 years, with the cutaneous, muscular and skeletal domains being the most affected (17). JDM-related damage was predicted by high disease activity and organ damage at 6 months post-diagnosis, suggesting that CYP are already set on a more severe JDM trajectory from the beginning of the disease, and that they may require more intense treatment and monitoring.

Despite the younger age of people affected by JSLE and JDM, several studies identified increased CVD-risk factors, such as endothelial dysfunction, increased arterial stiffness or presence of atherosclerotic lesions in paediatric populations (18, 19). The APPLE (Atherosclerosis Prevention in Paediatric Lupus Erythematosus) clinical trial tested the efficacy of atorvastatin treatment in JSLE (20), and although the trial did not meet the primary endpoint, the secondary analyses identified certain biomarkers and sub-cohort characteristics associated with increased CVD-risk and response to statin (21, 22). Clinicians remain concern about the lack of evidence-based strategies for management of CVD-risk in JSLE as highlighted by a recent international survey (10).

There are validated outcome measures of damage widely used both in research and in clinical practice in paediatric rheumatology, such as the paediatric SLE Damage Index (Ped-SDI)(23), similar to the validated tool used in adults with SLE (the SLICC Damage Index developed by the Systemic Lupus Collaborating Clinics), but which includes two additional items (puberty delay and growth retardation). Similarly, the Myositis Damage Index (MDI) evaluates the severity of disease-related damage across 11 domains on a visual analogue score (VAS) 1 to 10 and is validated for use across the life span. However, the assessment of CVD-risk in younger populations, and in particularly in the context of an underlying RMDs is more challenging (24). The CVD-risk scores validated for use in general population underperform in SLE, irrespective of the cohort age (9, 25), and they have not been tested before in JDM.

The most used CVD-risk scores in the general population are aimed at predicting CVD, including coronary death, myocardial infarction, coronary insufficiency, angina, ischemic stroke, haemorrhagic stroke, transient ischemic attack, peripheral artery disease, heart failure.

The Framingham Risk Score (FRS) estimates 10-year risk of CVD based on age, sex, total and high-density lipoprotein (HDL) cholesterol fractions, blood pressure, diabetes, and smoking status (26). A modified version (mFRS) has also been evaluated in adults with SLE, and found to misclassify 16.1% individuals grouped in the low-risk category by the presence of atherosclerotic plaque on vascular scans (25).

The atherosclerotic cardiovascular disease (ASCVD) risk calculator provided by the American Heart Association and the American College of Cardiology: estimates the 10-year and lifetime risks for CVD based on age, sex, race, total cholesterol, HDL cholesterol, systolic blood pressure, blood pressure lowering medication use, diabetes status, and smoking status (27). The model discriminates CVD risk poorly in younger individuals because age is the primary driver of the score. Consistent with this limitation, previous studies have also shown reduced performance in adults (25) and children with SLE (9).

QRISK3 is a 10-year or life-time model of CVD-risk estimation in general population that uniquely incorporates rheumatoid arthritis, SLE, and prolonged steroid use as risk enhancers (28), although misclassified 10% SLE adults with atherosclerosis plaque as low-risk in a recent study (25).

We are the first group to assess the performance of the PDAY (Pathobiological Determinants of Atherosclerosis in Adolescence) score in JSLE. Unlike the 10-year risk, the PDAY estimates “arterial age” or atherosclerotic burden based on risk factors. It was derived from autopsy studies of young accident victims and relates risk factors to actual arterial disease, and includes non-HDL and HDL cholesterol, smoking, blood pressure, body mass index (BMI), glucose, sex, and age (age-weighted, 1 point ≈ 1 year of vascular aging). Importantly, the PDAY focuses on cumulative risk burden, which makes it more sensitive for younger populations. Previous studies, such as CARDIA and Young Finns have shown that PDAY in adolescence predicted carotid intima-media thickness (CIMT) and coronary artery calcification years later. PDAY scores ≥1 or 2 identify adolescents with higher-than-normal clustering of risk factors, whereas higher thresholds (≥ 10) indicate increased CVD-risk, comparable to that of individuals 10 years older. PDAY adequately predicted atherosclerosis development early and later in life (29, 30).

Vascular scans to assess the presence of subclinical atherosclerosis are not routinely used in paediatric clinical practice, and therefore being able to assess the clinical utility of easy-to-implement CVD-risk scores has practical implications. Our group has previously found that FRS, ASCVD and QRISK3 failed to identify the majority of the high risk CYP with JSLE diagnosed based on CIMT progression over 3 years in the APPLE trial(9). Cumulatively, these adult CVD-risk scores appreciated that 86.3-100% CYP with JSLE had very low CVD-risk, while based on the CIMT progression in the placebo arm in the APPLE trial, 58.3% had high CVD-risk.

This study aimed to compare the CVD-risk estimation in young adults with JSLE and JDM using multiple scoring tools in these two populations.

The main hypothesis is that young adults with JSLE carry a higher CVD-risk burden than JDM and that the PDAY score would be more effective in risk stratifying this young, high-risk population than the traditional 10-year risk calculator.

This hypothesis was based on data derived from adult studies, such as the VITAL (Veterans With Premature Atherosclerosis) registry analysis, which identified SLE as the RMD associated with the highest likelihood (OR: 3.06, 95% CI: 2.38-3.93) of extremely premature atherosclerotic CVD (31). On the other hand, no significant differences in CVD-risk was found between adults with idiopathic inflammatory myopathies and age and sex-matched general population (32), suggesting that JDM may be associated with a lower CVD-risk that JSLE.

## Patients and methods

We conducted a single centre study with long-term follow-up, involving retrospective data collection to appreciate disease duration and age at disease onset in two cohorts of CYP and prospective data collection from the point of their transition to our adolescent rheumatology service through follow up into adulthood. All participants provided informed consent, and the study was approved by the London-Harrow Research Ethics Committee, reference 11/LO/0330. Demographic data, and data about disease manifestations and CVD-risk factors were collected cumulatively at the last assessment. Continuous variables were presented as mean ± standard deviation (SD) if normally distributed and as median (interquartile range, IQR) if skewed. Categorical variables were presented as counts and percentages. For key risk factors and scores, we compared JSLE with JDM using t-tests or Mann-Whitney U tests for continuous variables and Chi-Square test and the Z-test for proportions for categorical variables.

We assessed the distribution of each CVD-risk score in each disease group. We defined a high-risk category for each score based on adult guidelines. For example, FRS ≥10% 10-year risk = high risk in adults, 5-9.9% as intermediate risk, and <5% as low risk; QRISK3 ≥10% and ASCVD >7.5% as high risk, as per accepted cut-offs. For PDAY, we considered >8 points as moderate to high risk due to the lack of established cutoffs. We examined the associations of damage and CVD-risk using multivariable logistic regression accounting for relevant variables, such as age, disease duration, BMI, low density lipoprotein (LDL)-cholesterol serum levels, and cumulative doses or duration of various treatments, and disease activity, using validated outcomes: e.g. presence of BILAG A/2 BILAG B episodes or SLEDAI score≥6 defined active JSLE, while MMT8 (the manual muscle testing including 8 muscle groups); CDASI (Cutaneous Dermatomyositis Disease Area and Severity Index) and serum muscle enzymes were used to appreciate JDM activity. Sensitivity analysis: We analyzed comparatively groups stratified based on damage and CVD-risk. Two-tailed p-value < 0.05 was considered statistically significant.

## Results

The mean age of the JSLE cohort (n=76) at the last follow-up was 24.3 years (SD 4.2 years), which was significantly older than that of the JDM cohort (n=79, mean age 20.1 ± 5.0 years, p<0.001), although there was no significant difference in the disease duration as usually JDM is diagnosed at a younger age than non-monogenic SLE, which starts around puberty. There was a significant female predominance in both cohorts (85% vs. 72% in the JSLE vs. JDM cohort, respectively) reflecting the well-recognized female bias in autoimmunity, and a significant longer delay from symptoms onset to diagnosis in JSLE compared to JDM (median 1.2 vs. 0.4 years, respectively) (**Table 1**). Both disease groups had adequate ethnic representation, reflecting the UK population diversity in London and neighboring geographical areas the cohorts were recruited from, and known trends in the prevalence of JSLE (more commonly affecting Asian and Black children) and JDM (more frequently encountered in White children).

**Table 1:**
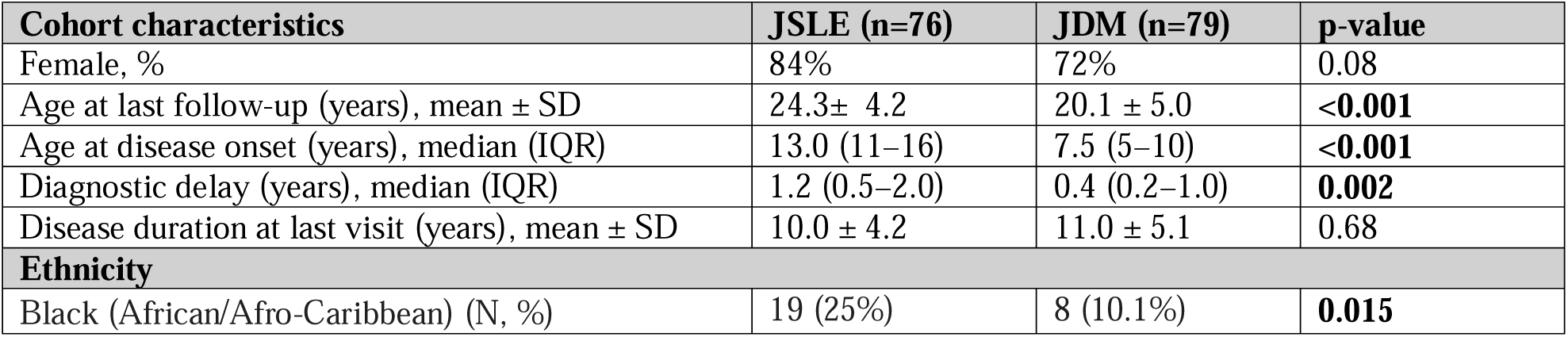
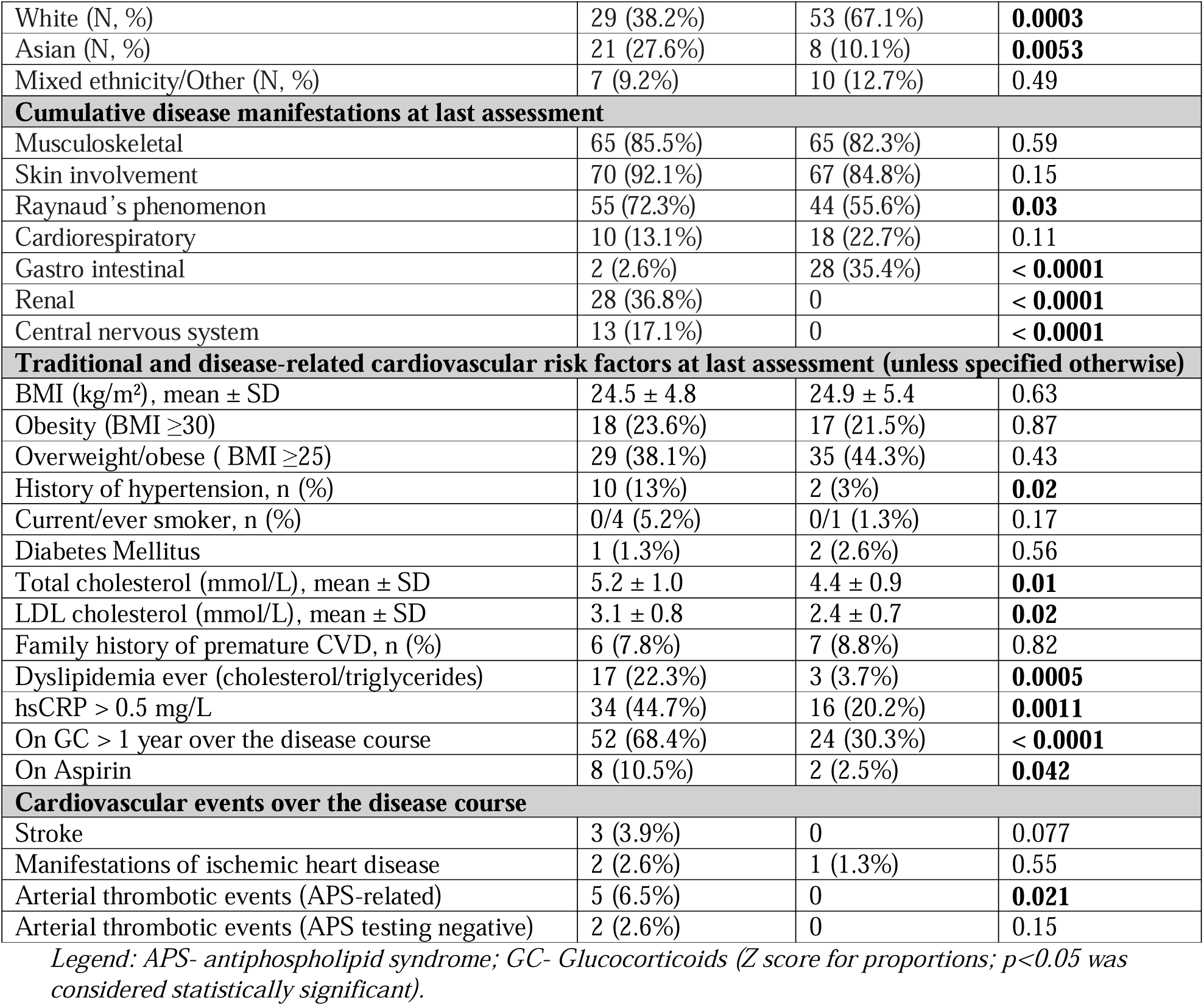
Cohort characteristics and prevalence of organ manifestations and traditional cardiovascular risk factors at the last assessment.

| Cohort characteristics | JSLE (n=76) | JDM (n=79) | p-value |
| --- | --- | --- | --- |
| Female, % | 84% | 72% | 0.08 |
| Age at last follow-up (years), mean ± SD | 24.3 ± 4.2 | 20.1 ± 5.0 | **<0.001** |
| Age at disease onset (years), median (IQR) | 13.0 (11–16) | 7.5 (5–10) | **<0.001** |
| Diagnostic delay (years), median (IQR) | 1.2 (0.5–2.0) | 0.4 (0.2–1.0) | **0.002** |
| Disease duration at last visit (years), mean ± SD | 10.0 ± 4.2 | 11.0 ± 5.1 | 0.68 |
| **Ethnicity** |  |  |  |
| Black (African/Afro-Caribbean) (N, %) | 19 (25%) | 8 (10.1%) | **0.015** |
| White (N, %) | 29 (38.2%) | 53 (67.1%) | **0.0003** |
| Asian (N, %) | 21 (27.6%) | 8 (10.1%) | **0.0053** |
| Mixed ethnicity/Other (N, %) | 7 (9.2%) | 10 (12.7%) | 0.49 |
| **Cumulative disease manifestations at last assessment** |  |  |  |
| Musculoskeletal | 65 (85.5%) | 65 (82.3%) | 0.59 |
| Skin involvement | 70 (92.1%) | 67 (84.8%) | 0.15 |
| Raynaud's phenomenon | 55 (72.3%) | 44 (55.6%) | **0.03** |
| Cardiorespiratory | 10 (13.1%) | 18 (22.7%) | 0.11 |
| Gastro intestinal | 2 (2.6%) | 28 (35.4%) | **< 0.0001** |
| Renal | 28 (36.8%) | 0 | **< 0.0001** |
| Central nervous system | 13 (17.1%) | 0 | **< 0.0001** |
| **Traditional and disease-related cardiovascular risk factors at last assessment (unless specified otherwise)** |  |  |  |
| BMI (kg/m<sup>2</sup>), mean ± SD | 24.5 ± 4.8 | 24.9 ± 5.4 | 0.63 |
| Obesity (BMI ≥30) | 18 (23.6%) | 17 (21.5%) | 0.87 |
| Overweight/obese ( BMI ≥25) | 29 (38.1%) | 35 (44.3%) | 0.43 |
| History of hypertension, n (%) | 10 (13%) | 2 (3%) | **0.02** |
| Current/ever smoker, n (%) | 0/4 (5.2%) | 0/1 (1.3%) | 0.17 |
| Diabetes Mellitus | 1 (1.3%) | 2 (2.6%) | 0.56 |
| Total cholesterol (mmol/L), mean ± SD | 5.2 ± 1.0 | 4.4 ± 0.9 | **0.01** |
| LDL cholesterol (mmol/L), mean ± SD | 3.1 ± 0.8 | 2.4 ± 0.7 | **0.02** |
| Family history of premature CVD, n (%) | 6 (7.8%) | 7 (8.8%) | 0.82 |
| Dyslipidemia ever (cholesterol/triglycerides) | 17 (22.3%) | 3 (3.7%) | **0.0005** |
| hsCRP > 0.5 mg/L | 34 (44.7%) | 16 (20.2%) | **0.0011** |
| On GC > 1 year over the disease course | 52 (68.4%) | 24 (30.3%) | **< 0.0001** |
| On Aspirin | 8 (10.5%) | 2 (2.5%) | **0.042** |
| **Cardiovascular events over the disease course** |  |  |  |
| Stroke | 3 (3.9%) | 0 | 0.077 |
| Manifestations of ischemic heart disease | 2 (2.6%) | 1 (1.3%) | 0.55 |
| Arterial thrombotic events (APS-related) | 5 (6.5%) | 0 | **0.021** |
| Arterial thrombotic events (APS testing negative) | 2 (2.6%) | 0 | 0.15 |

Legend: APS- antiphospholipid syndrome; GC- Glucocorticoids (Z score for proportions; $p < 0.05$ was considered statistically significant).

In terms of organ involvement, gastro-intestinal (G-I), renal and central nervous system (CNS) involvement, as well as Raynaud’s’ phenomenon, were more common in JSLE.

### Assessment of the prevalence of traditional CVD-risk factors at last assessment

AYA with JSLE have significantly increased proportion of individuals with traditional CVD-risk factors, including history of hypertension and dyslipidemia; they also had a higher serum total cholesterol and LDL-cholesterol levels at the last assessment compared to AYA with JDM (**Table 1**). Despite no significant differences between the two cohorts, a higher proportion of AYA with JSLE were overweight or obese at the last assessment (38.1 % vs. 44.3%, respectively, p=0.43).

Cumulatively 13/76 (17.1%) and 3/76 (3.9%) AYA with JSLE have been treated with antihypertensive treatment or statins, respectively, during episodes of acute lupus nephritis (LN), while none of the AYA with JDM received such treatments. At last visit, 44.7% of JSLE vs. 20.2% of JDM cohort had an increased high sensitivity C-Reactive Protein (hsCRP) level (p=0.0011), which was not related to disease flares or concomitant infections, which could reflect subclinical low degree inflammation with relevance for the CVD-risk. Importantly, a higher proportion of AYA with JSLE have been treated with GC cumulatively for longer than 1 year over their disease course (68.4% vs. 30. 3%, p< 0.0001).

Despite the relatively low proportion of AYA who experienced CVD events throughout their disease course, almost all of them were diagnosed with JSLE with or without concomitant antiphospholipid syndrome (APS) (**Table 1**).

### Treatments used in both cohorts throughout the disease course

As the increased CVD-risk seen in AYA with RMD is in high proportion attributable to the disease-driven chronic inflammation, to the impact of treatment as well as that of traditional CVD-risk factors, we also assessed comparatively all the medications used in both cohorts throughout the disease course. As expected, a higher proportion of AYA with JSLE than those with JDM have been treated or were currently treated with GC (p=0.07), B cell depletion therapy (p=0.00006), mycophenolate mofetil (p=0.0015) or hydroxychloroquine (p=0.00001), while for the majority of other therapies, including the ones used only in a minority of individuals, there were no statistically significant differences between the JSLE and JDM cohorts (**Table 2**).

**Table 2:**
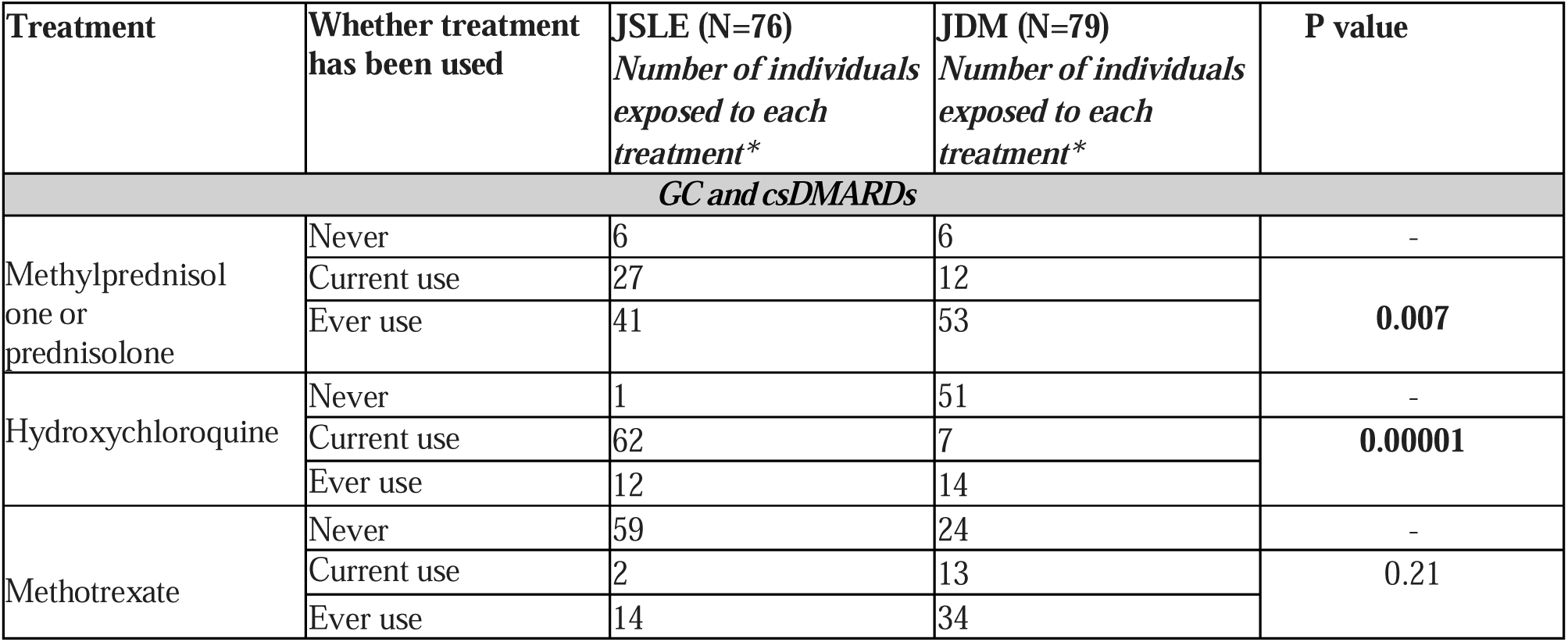
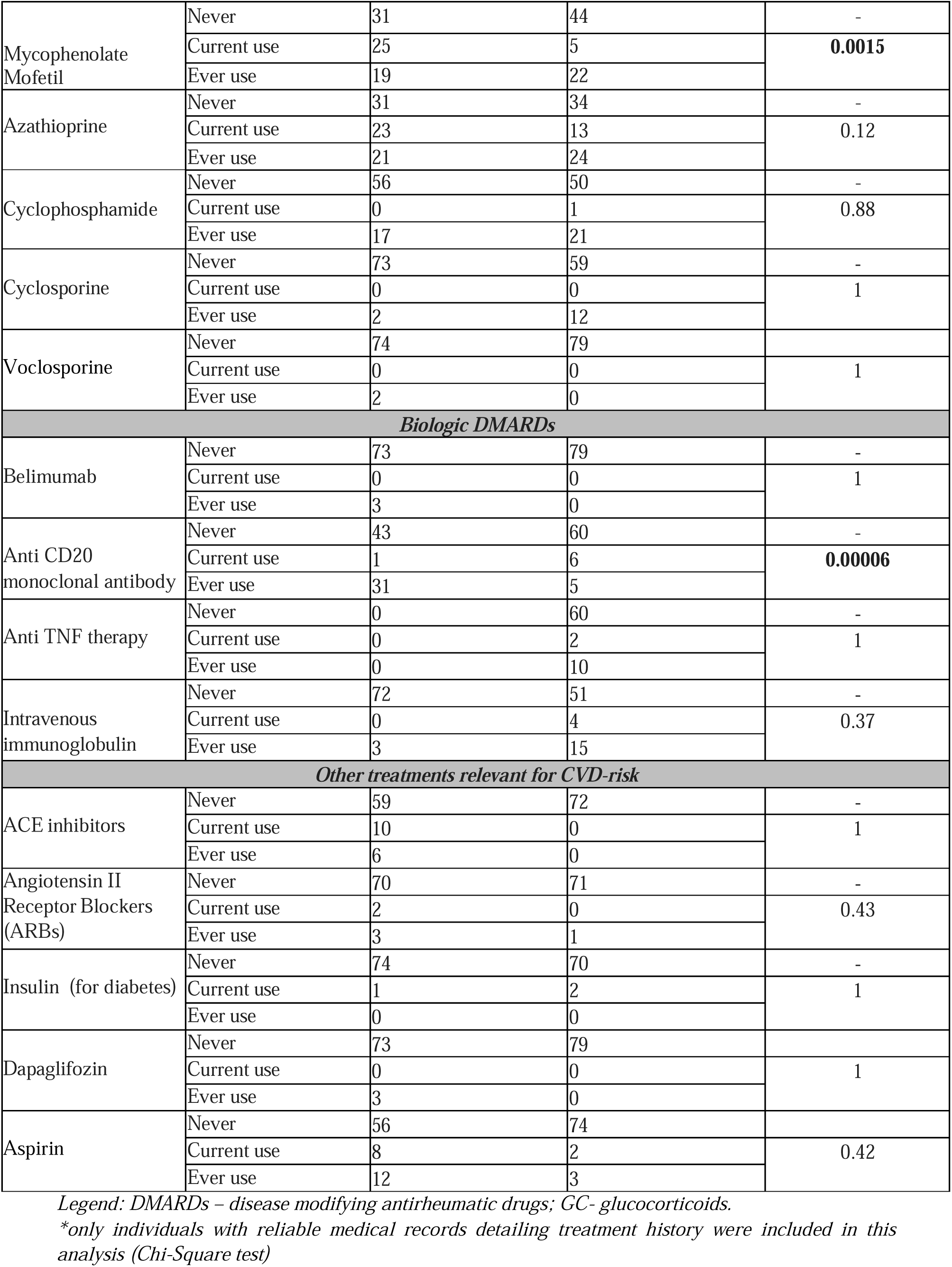
Treatment exposures throughout the disease course in the JSLE compared to the JDM cohort.

### Assessment of damage in JSLE vs. JDM

We assessed comparatively the damage accrual across both cohorts and across various disease domains. Overall, 39 (51.3%) AYA with JSLE accumulated damage over the disease course vs. vs. 47 (59.4%) AYA with JDM (p=0.307).

A higher proportion of AYA with JDM accumulated musculoskeletal, cutaneous, pulmonary and G-I damage compared to JSLE, while peripheral vascular, renal, and CNS domains were more commonly involved in JSLE. Additionally, AYA with JSLE had increased malignancy and mortality overall over the disease course, with two individuals having died, both at ages 20-25 years, from a combination of severe JSLE flares and infective complications (**Table 3**).

**Table 3:** Comparative assessment of damage accrual in JSLE vs. JDM over the disease course.

| Affected organs and systems | AYA with JSLE (N=76) and damage (N, %) | AYA with JDM (N=79) and damage (N, %) | P value |
| --- | --- | --- | --- |
| Damage in at least one domain | 39 (51.3%) | 47 (59.4%) | 0.307 |
| Musculoskeletal system | 2 (2.7%) | 30 (39.4%) | **< 0.0001** |
| Cutaneous | 9 (12%) | 39 (49.3%) | **< 0.0001** |
| Gastrointestinal | 2 (2.7%) | 13 (16.4%) | **0.004** |
| Pulmonary | 1 (1.3%) | 9 (11.3%) | **0.011** |
| Cardiovascular | 6 (8%) | 8 (10.1%) | 0.64 |
| Peripheral vascular | 5 (6.3%) | 2 (2.5%) | 0.21 |
| Endocrine (delayed puberty) | 1 (1.3%) | 6 (7.5%) | 0.06 |
| Ocular | 1 (1.3%) | 2 (2.5%) | 0.58 |
| Malignancy | 3 (4%) | 0 | 0.07 |
| Neuropsychiatric | 9 (12%) | 0 | **0.0015** |
| Renal | 8 (10.6%) | 0 | **0.0028** |
| Death | 2 (2.7%) | 0 | 0.14 |

*Legend: AYA- adolescents and young adults; JDM – juvenile dermatomyositis; JSLE – juvenile systemic lupus erythematosus.*

In the JSLE cohort, AYA who accrued damage had increased use of oral prednisolone (p=0.043) and cyclophosphamide (p=0.01), as well as had in a lower proportion a median BILAG score =0 and in a higher proportion median BILAG scores of 1-8 and >8 throughout their follow-up (p=0.04) (**Table 4**). In a multivariable regression analysis, the SDI score was independently predicted by the cumulative cyclophosphamide dose (SDI score increased by 1 point for each 1.6g of cyclophosphamide, p = 0.043), and BMI (SDI increased by 1 point for each 3.43 kg/m^2^ increase in BMI, p = 0.038), AYA with JDM associated with damage were slightly older age at diagnosis (p=0.003), had slightly longer disease duration (p=0.006) and delay in diagnosis (p=0.052), higher BMI (p=0.04) and patient global VAS (<0.001) than the ones without damage (**Table 4**). Additionally, AYA with JDM-associated damage were treated in a higher proportion with hydroxychloroquine (30/47 vs. 7/32, p=0.022) throughout their disease course compared to AYA without damage, while there were no differences in other treatments, including cumulative dose of GC (p=0.279). In a multivariable regression analysis, the MDI was independently predicted only by BMI (MDI increased by 1 point for each 1.039 kg/m^2^ increase in BMI, p=0.026) and by the physician damage VAS assessment, which is not surprising as both outcomes are reported by physicians (MDI increased by one point for each 1.166 point increase in physician VAS score on a scale 0-10, p=0.027).

**Table 4:**
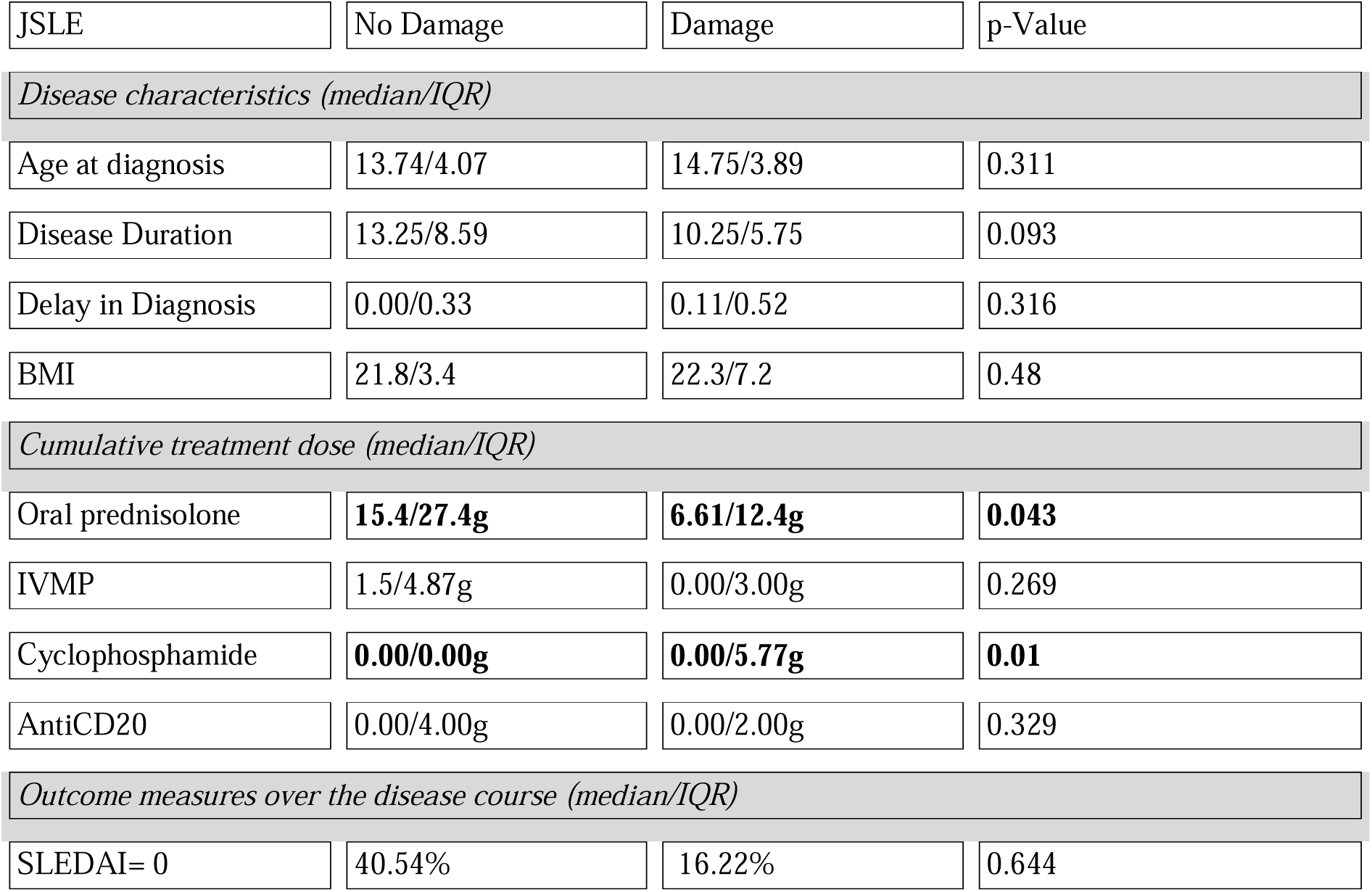
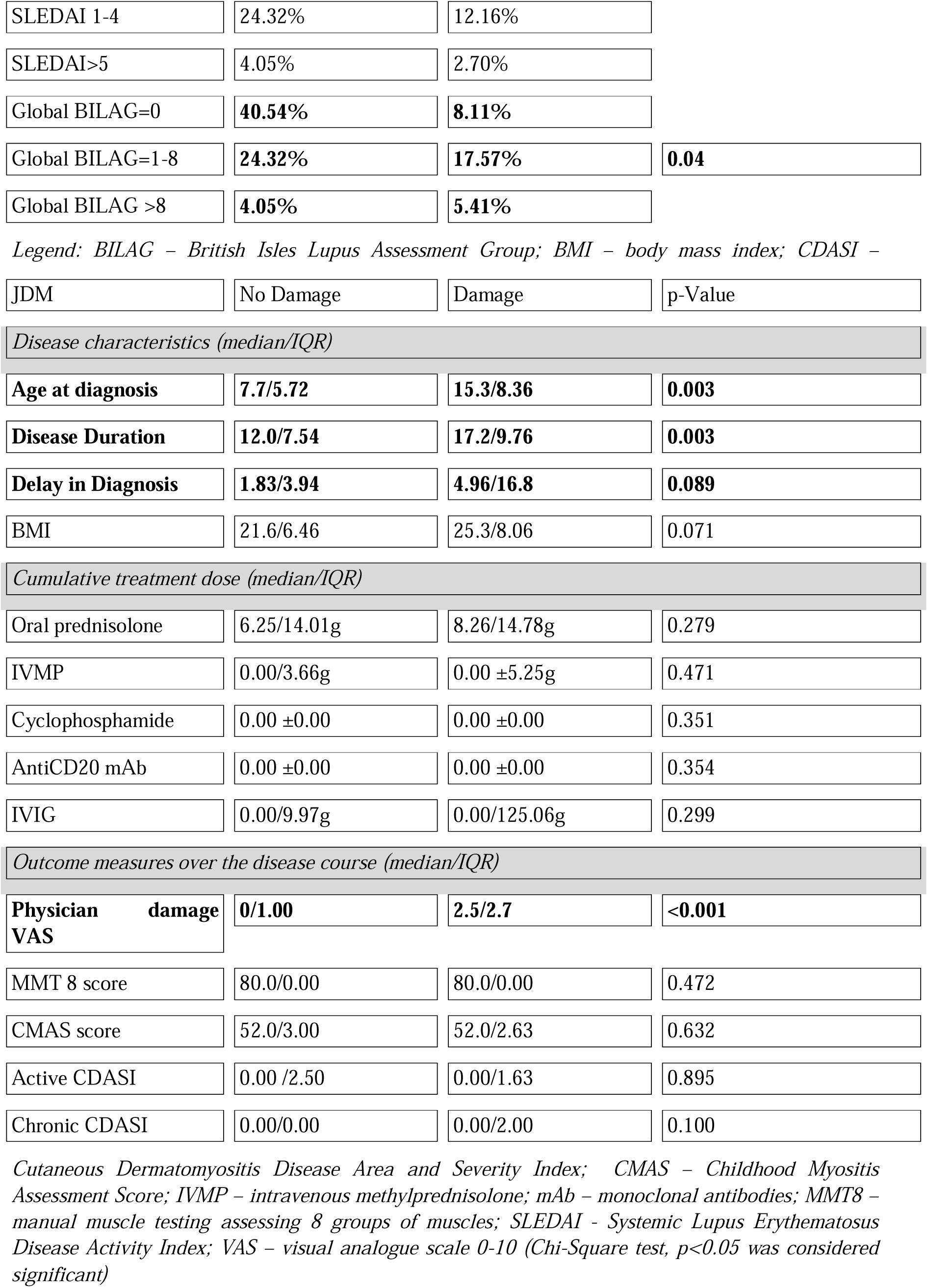
Comparative assessment of JSLE and JDM sub-cohorts with damage vs. no damage.

| JSLE | No Damage | Damage | p-Value |
| --- | --- | --- | --- |
| *Disease characteristics (median/IQR)* |  |  |  |
| Age at diagnosis | 13.74/4.07 | 14.75/3.89 | 0.311 |
| Disease Duration | 13.25/8.59 | 10.25/5.75 | 0.093 |
| Delay in Diagnosis | 0.00/0.33 | 0.11/0.52 | 0.316 |
| BMI | 21.8/3.4 | 22.3/7.2 | 0.48 |
| *Cumulative treatment dose (median/IQR)* |  |  |  |
| Oral prednisolone | **15.4/27.4g** | **6.61/12.4g** | **0.043** |
| IVMP | 1.5/4.87g | 0.00/3.00g | 0.269 |
| Cyclophosphamide | **0.00/0.00g** | **0.00/5.77g** | **0.01** |
| AntiCD20 | 0.00/4.00g | 0.00/2.00g | 0.329 |
| *Outcome measures over the disease course (median/IQR)* |  |  |  |
| SLEDAI= 0 | 40.54% | 16.22% | 0.644 |
| SLEDAI 1-4 | 24.32% | 12.16% |  |
| SLEDAI>5 | 4.05% | 2.70% |  |
| Global BILAG=0 | **40.54%** | **8.11%** |  |
| Global BILAG=1-8 | **24.32%** | **17.57%** | **0.04** |
| Global BILAG >8 | **4.05%** | **5.41%** |  |

Legend: BILAG – British Isles Lupus Assessment Group; BMI – body mass index; CDASI –
| JDM | No Damage | Damage | p-Value |
| --- | --- | --- | --- |
| Disease characteristics (median/IQR) |  |  |  |
| Age at diagnosis | 7.7/5.72 | 15.3/8.36 | 0.003 |
| Disease Duration | 12.0/7.54 | 17.2/9.76 | 0.003 |
| Delay in Diagnosis | 1.83/3.94 | 4.96/16.8 | 0.089 |
| BMI | 21.6/6.46 | 25.3/8.06 | 0.071 |
| Cumulative treatment dose (median/IQR) |  |  |  |
| Oral prednisolone | 6.25/14.01g | 8.26/14.78g | 0.279 |
| IVMP | 0.00/3.66g | 0.00 ±5.25g | 0.471 |
| Cyclophosphamide | 0.00 ±0.00 | 0.00 ±0.00 | 0.351 |
| AntiCD20 mAb | 0.00 ±0.00 | 0.00 ±0.00 | 0.354 |
| IVIG | 0.00/9.97g | 0.00/125.06g | 0.299 |
| Outcome measures over the disease course (median/IQR) |  |  |  |
| Physician damage VAS | 0/1.00 | 2.5/2.7 | <0.001 |
| MMT 8 score | 80.0/0.00 | 80.0/0.00 | 0.472 |
| CMAS score | 52.0/3.00 | 52.0/2.63 | 0.632 |
| Active CDASI | 0.00 /2.50 | 0.00/1.63 | 0.895 |
| Chronic CDASI | 0.00/0.00 | 0.00/2.00 | 0.100 |

Cutaneous Dermatomyositis Disease Area and Severity Index; CMAS – Childhood Myositis Assessment Score; IVMP – intravenous methylprednisolone; mAb – monoclonal antibodies; MMT8 – manual muscle testing assessing 8 groups of muscles; SLEDAI - Systemic Lupus Erythematosus Disease Activity Index; VAS – visual analogue scale 0-10 (Chi-Square test, $p<0.05$ was considered significant)

### Assessment of CVD-risk using validated adult scores

Despite no disease specific recommendations for assessment of CVD-risk in JSLE or JDM in adulthood exist, we stratified both cohorts based on the categories of risk defined by the most commonly used CVD-risk tools used in the general population: the FRS and ASCVD risk scores, in addition to the QRISK3 risk score, which is the only one accounting for a diagnosis of SLE and GC treatment use as independent risk factors. The JSLE cohort had significantly higher FRS and ASCVD scores than the JDM cohort (**Figure 1**), but absolute values were low for both (medians 0–0.5%). QRISK3 yielded higher risk estimates in JSLE compared to JDM (median 2.3%, IQR =1–5% vs. median 1%, IQR = 0-4%, P= 0.048). Notably, none of AYA with JSLE or JDM reached the conventional moderate CVD-risk threshold of ≥10% as assessed by the FRS and QRISK3 scores, while only one adult with JSLE had a moderate CVD-risk as estimated by a ASCVD risk score of ≥ 7.5% (all adult CVD-risk scores estimate the 10-year risk of a CVD-related event).

**Figure 1:**
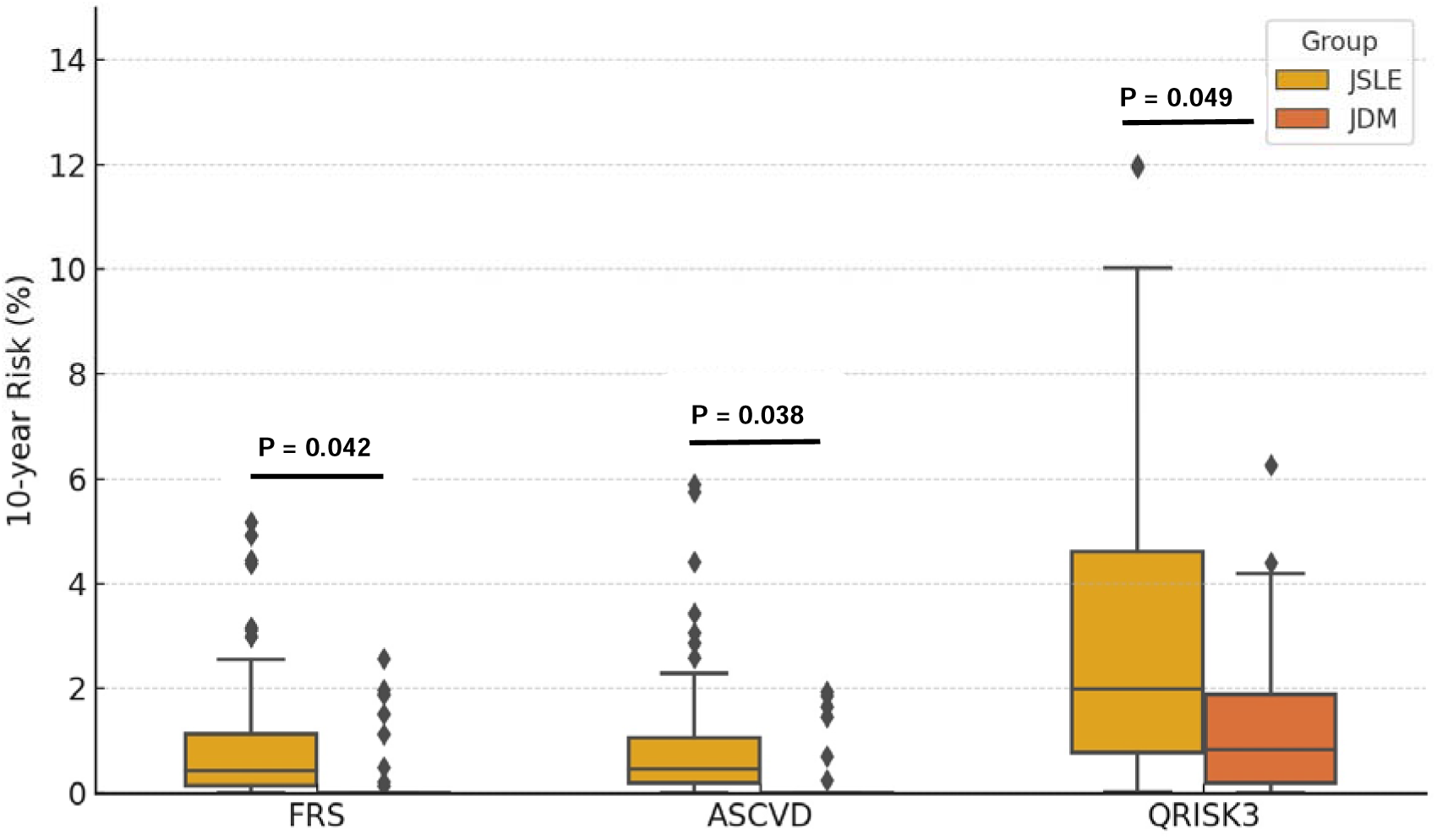
Boxplots show median and interquartile range for the adult CVD-risk scores, with individual risk values (% at 10 years) overlaid Legend: ASCVD - atherosclerotic cardiovascular disease risk score; FRS – Framingham Risk Score; JDM – juvenile dermatomyositis; JSLE – juvenile systemic lupus erythematosus (Mann-Whitney U test, P<0.05 was considered significant).

### Assessment of CVD-risk using a validated paediatric score (PDAY)

In the JSLE cohort, the PDAY score ranged widely: from -2 (in individuals with best features) to 23 points (reflecting additional 23 years of vascular ageing). PDAY score stratified the JSLE cohort as following: 18.4% had low risk (0 or less points), 22.3% had moderate risk (1-5 points), 25 % had high (6-10 points), and 34.2% had very high (>10 points) CVD-risk. The median PDAY for the JSLE cohort was 5 points (IQR 4-13), with a mean of 7.8. Notably, 43.5% of AYA with JSLE had a PDAY ≥8 (the defined cut-off for high and very high CVD-risk), and 32.8% had a PDAY >10 (very high CVD-risk).

In contrast, in the JDM cohort, the PDAY score was significantly lower (median 0, IQR 0-3, p=0.0001). Only 12 (15.1%) AYA with JDM had a PDAY ≥ 8, most of whom were male adults with ongoing JDM inflammation at last assessment, while 50% cohort had a PDAY score = 0 or less, highlighting very good vascular health.

### Predictors of high and very high CVD-risk in JSLE

We further evaluated the predictors of PDAY ≥ 8 points in the JSLE sub-cohort stratified as high and very high CVR-risk (N=33, 43.5% of JSLE patients) using a multivariable logistic model. Four independent predictors were found: older age (OR ∼1.5 per 5-year increase in age, p=0.03), longer disease duration (OR ∼1.4 per 5-year increase, p=0.04), higher LDL level (OR ∼2.0 per 1 mmol/L increase, p=0.02), and presence of active lupus (OR ∼3.3 for any flare defined as either one BILAG A or two BILAG B scores or SLEDAI ≥6 at last visit, p=0.01), when the exposure to GC for longer than 1 year without interruption and the BMI measurements were included in the model. Despite being recognized as a main driver of CVD-risk, the duration of treatment with GC has not been shown to be an independent factor for high CVD-risk in JSLE in our sub-cohort, although exposure to GC may contribute to worse lipid profile as well as reflect more active disease, factors which both independently contributed to a PDAY score ≥ 8 (**Table 5**).

**Table 5.** Multivariable logistic regression analysis of predictors for moderate-high CVD-risk (PDAY ≥ 8) in the JSLE cohort (N=33).

| Predictor | Odds Ratio (95% CI) | p-value |
| --- | --- | --- |
| Age (per 5-year increase) | 1.5 (1.1 - 2.1) | 0.03 |
| Disease duration (per 5-year) | 1.4 (1.0 - 2.0) | 0.04 |
| LDL cholesterol (per 1 mmol/L) | 2.0 (1.1 - 3.6) | 0.02 |
| Active disease at last visit | 3.3 (1.3 - 8.4) | 0.01 |
| BMI (per 5 kg/m<sup>2</sup>) | 1.2 (0.8 - 1.8) | 0.33 |
| Duration of steroid use >1yr | 1.5 (0.5 - 4.4) | 0.45 |

*Legend: BMI – body mass index; CI – confidence interval; LDL – low density lipoprotein. Active disease at last visit was defined as one BILAG A or two BILAG B scores or SLEDAI 2K $\geq 6$ ( $p < 0.05$ was considered significant)*

A similar robust modelling could not be undertaken in the JDM cohort, because of the small number of AYA with high PDAY score (n=12). However, we evaluated the individuals with JDM with high PDAY scores, and they were all males, one was an ex-smoker (+2 points), and all 12 had either low HDL and/or active disease at the last assessment when their PDAY score was evaluated. Interestingly, none of the parameters associated with active JDM (increase in muscle enzyme levels, low MMT 8 score or the presence of active skin disease assessed by the active CDASI score) showed an association with PDAY, providing some evidence that traditional CVD-risk were likely the drivers of the increase in the PDAY score in the JDM cohort.

### Atherosclerosis detected by vascular investigations in individuals with high PDAY scores

Although routine vascular screening for CVD-risk evaluation is not embedded in clinical practice, we had the opportunity to evaluate the performance of PDAY score against imaging evidence for atherosclerosis in five AYA with JSLE and two with JDM. All five AYA with JSLE had increased CIMT measurements above the age-range on carotid artery ultrasound and PDAY scores ≥10. Interestingly, none of them had increased CVD-risk based on FRS, ASCVD or QRISK 3 scores (all were close to 0%). Two AYA with JDM had abnormal brachial flow mediated dilation (FMD) results, indicating endothelial dysfunction, measured using the EndoPAT device, which evaluates the reactive hyperemic index (RHI), which was ∼1.5 in both cases, below the normal cutoff of 1.67. Both AYA with JDM had a PDAY score =10, while their adult CVD-risk scores were 0. This provides evidence of a good correlation of PDAY scores with the presence of atherosclerosis or endothelial dysfunction in a small sub-cohort of AYA with JSLE and JDM.

### Predictors of CVD-risk assessed using the PDAY score in both cohorts

When we compared the JSLE vs JDM cohort, the PDAY scores remained significantly higher in JSLE even after adjusting for age differences (p<0.001) as the JSLE cohort was slightly older. In an age-matched comparison (looking only at individuals aged 18-22 years in both cohorts), PDAY also remained higher in JSLE (median 5 vs 0, p<0.001).

In the JSLE cohort, AYA with active disease (SLEDAI≥6 or one BILAG A or two BILAG B scores) at last visit had a mean PDAY of 11, compared to 7 for those with inactive disease (p=0.004). AYA with JSLE and organ damage (SDI ≥1) also tended to have higher PDAY scores (mean 10 vs 6, p=0.06). Interestingly, serological tests (e.g. anti-dsDNA or C3 and C4 complement levels) at the last assessment did not directly correlate with the PDAY score. In the JDM cohort, the PDAY score was not influenced by disease activity or other parameters either (with a caveat of a small sample size as only 12 AYA with JDM had high PDAY scores).

Despite SLE being an independent risk classifier for QRISK3 score, its ability to capture risk in this JSLE cohort was limited, but higher than that of FRS (median 0.2% vs. 0.0%, p<0.01). For example, a 25-year-old with JSLE on GC and with a high BMI has only a ∼2% risk of a cardiovascular event in the next 10 years, as estimated by the QRISK3-score, which explains why this entire JSLE cohort had very low scores overall.

## Conclusion

This study is a real-life investigation of long-term outcomes in a large single-centre cohort of AYA with JSLE and JDM focused on damage accrual and assessment of CVD-risk using validated tools and data routinely collected in clinical practice.

The results highlighted differences in long-term outcomes between JSLE and JDM, conditions usually looked after in rheumatology services specialized in delivering multi-disciplinary care for individuals with a diagnosis of connective tissue diseases. This study integrated electronic health records data from paediatric to adolescent rheumatology services (supported by a robust transition process) and from adolescent to a young adult rheumatology service delivered by the same clinical team and led by a consultant rheumatologist trained across the age span, interested in objectively assessing the disease activity and damage as well as implementing treat to target strategies at each routine appointment, which facilitated data collection. The damage accrual observed across organs and systems in this JSLE cohort is similar to than that observed in the large UK JSLE cohort in the first year post diagnosis (51.3% vs. 54.4%, despite our cohort having a significantly longer follow-up (10 years vs. 3.6 years in the UK JSLE cohort)(33), which may reflect the potential added benefit of active implementation of treat to target strategies in our clinical service (34). We also observed a lower prevalence of damage in our JDM cohort when compared to a historical cohort of similar size (59.4% vs. 90%)(17) or with a contemporaneous larger JDM cohort (where damage has been observed in 95% cases after a median of 21.8 years followed up)(35). In the last study, 53% JDM cohort had active disease, 21.8 years post-diagnosis, while in our cohort, only 15.1% had persistently active disease at the last assessment, which may explain the disparity in the disease activity and damage trajectories in these two cohorts. We found that damage accrual in both JSLE and JDM was independently predicted by BMI, in addition to cumulative cyclophosphamide dose for JSLE, and physician damage VAS for JDM, as assessed using multivariable regression models. AYA with damage has increased disease activity throughout the disease course in both cohorts, highlighting the need for tighter disease control to prevent damage in both JSLE and JDM and need for objective assessment and aim to achieve low disease activity states (36, 37).

Similarly to previous observations in JSLE and adults with SLE, we also found that adult CVD-risk scores validated for use in general populations do not reliably identify individuals with SLE at risk, and have no clinical utility (24, 25). We previously tested the performance of PDAY in a combined cohort of children from the USA (recruited to the APPLE trial) and AYA with JSLE recruited from University College London and found better performance of PDAY in young adults compared to children with JSLE when assessed against a metabolomic signature of atherosclerosis progression or evaluation of CIMT progression on vascular scans (9), providing additional evidence that this CVD-risk score may have some potential clinical utility in JSLE. Overall, all CVD-risk scores showed increased estimated CVD-risk in JSLE compared to JDM, even after adjusting for age and disease duration. This real-life longitudinal cohort evaluation also showed less CVD and decreased contribution of disease activity in driving CVD-risk in JDM compared to JSLE, which has not been investigated before.

First, due to the partly retrospective data collection and the single-centre nature of the study, our findings may reflect specific management practices and may not be universally generalizable. Second, due to the young age of both cohorts and low incidence of cardiovascular events, and lack of vascular imaging for assessment of atherosclerosis (except for a small sub-cohort), the predictive accuracy of PDAY score has been inferred rather than directly confirmed.

Despite this, the adequate sample size balanced across both diseases and mean disease duration of over 10 years, provides the first comparative insight into the long-term outcomes and CVD-risk assessment of JSLE and JDM in young adulthood, relevant for resource allocation to support clinical services in which these individuals are followed-up.

In terms of clinical relevance, these findings strongly support the argument for using AYA-specific tools for CVD-risk assessment, such as the PDAY score, which is very easy to use, and may have some benefits in capturing AYA with JSLE at risk, as well as highlight that further research into better biomarkers for robust CVD-risk stratification (2, 10).

Sole reliance on traditional CVD-risk scores or management of this risk in primary care may significantly underestimate this risk in AYA with RMDs and delay the implementation of preventive interventions. In conjunction with assessments such as the PDAY score, clinicians should maintain a high level of vigilance for identification and investigation of individuals at risk, as well as implement tighter disease control, and minimize the exposure to GC, allowing for timely initiation of lifestyle modifications, risk factors monitoring, and potentially pharmacological interventions in selected categories of AYA with JSLE.

In conclusion, this study demonstrates that traditional short-term CVD-risk assessment tools have limited utility in AYA RMDs, and highlights differences in damage accrual and long-term CVD-risk in JSLE vs. JDM. Our study adds weight to calls for specific CVD-risk stratification tools and tailored interventions for younger populations with RMDs. Ultimately, improving outcomes in JSLE and JDM will require a holistic approach that includes not only better disease control, but also safeguarding of cardiovascular health decades from the disease onset.

Future longitudinal, multicenter studies with good geographic and ethnic representability are recommended to validate these findings further and establish the potential clinical utility of PDAY or other similar AYA-specific assessment tools for CVD-risk stratification in rheumatology practice.

## Ethics statement

The study has been approved by the London-Harrow Research Ethics Committee, reference 11/LO/0330. All participants have provided written consent.

## Contributorship

CC conceptualised the study and generated data related to outcome measures and CVD-risk assessment. JL, IA, TM, SD, NR, EP collected clinical and laboratory data under the supervision of CC. JL, IA, TM, SD, NR, EP and CC analysed the data. CC wrote the manuscript. All authors critically appraised the manuscript and approved the final version.

## Data availability

The data used in this study will be made available upon reasonable request to the corresponding author, subject to approval from the study steering committee and completion of a data transfer agreement.

No AI has been used in the writing of this manuscript beyond checking for spelling errors.

